# Association of alcohol consumption and frequency with loneliness: A cross-sectional study among Japanese workers during the COVID-19 pandemic

**DOI:** 10.1101/2021.11.05.21265962

**Authors:** Yusuke Konno, Makoto Okawara, Ayako Hino, Tomohisa Nagata, Keiji Muramatsu, Seiichiro Tateishi, Mayumi Tsuji, Akira Ogami, Reiji Yoshimura, Yoshihisa Fujino, for the CORoNaWork Project

**Affiliations:** Department of Environmental Epidemiology, Institute of Industrial Ecological Sciences, University of Occupational and Environmental Health, Japan, Kitakyushu, Japan; Department of Psychiatry, University of Occupational and Environmental Health, Japan, Kitakyushu, Japan; Department of Mental Health, Institute of Industrial Ecological Sciences, University of Occupational and Environmental Health, Japan, Kitakyushu, Japan; Department of Occupational Health Practice and Management, Institute of Industrial Ecological Sciences, University of Occupational and Environmental Health, Japan, Kitakyushu, Japan; Department of Preventive Medicine and Community Health, School of Medicine, University of Occupational and Environmental Health, Japan, Kitakyushu, Japan; Department of Occupational Medicine, School of Medicine, University of Occupational and Environmental Health, Japan, Kitakyushu, Japan; Department of Environmental Health, School of Medicine, University of Occupational and Environmental Health, Japan, Kitakyushu, Japan; Department of Work Systems and Health, Institute of Industrial Ecological Sciences, University of Occupational and Environmental Health, Japan, Kitakyushu, Japan

## Abstract

**Background:** There are increasing concerns that prevention measures against coronavirus disease 2019 (COVID-19) such as social distancing and telework are leading to loneliness and poor lifestyle habits like increased alcohol consumption. The purpose of this study was to assess whether loneliness reported among workers during the COVID-19 pandemic is associated with changes in alcohol consumption.

**Methods:** The study comprised a cross-sectional, online survey of 27,036 workers between December 22 and 26, 2020. A questionnaire was used to assess loneliness, usual alcohol consumption and whether that consumption had changed. The odds ratios (ORs) were estimated by logistic regression analysis.

**Results:** A total of 2831 (10.5%) workers indicated they had increased alcohol consumption during the pandemic. Increased alcohol consumption was significantly associated with loneliness (OR=1.94, 95%CI 1.70–2.21). This association held true for those who indicated they were drinking two or more days per week (OR=1.98 95%CI 1.71–2.30) and those who drank less than one day per week (OR=1.51 95%CI 0.71–3.25). In contrast, there was no association between increased drinking and loneliness among those who indicated they hardly ever drank (OR=1.22 95%CI 0.55–2.72).

**Conclusions:** Among those with a drinking habit, increased alcohol consumption is associated with loneliness.

## Introduction

The first reported cases of coronavirus disease 2019 (COVID-19) emerged in December 2019 from Wuhan, China. Thereafter, the infection caused by SARS-CoV-2 spread worldwide and the situation was declared a pandemic by the World Health Organization (WHO) on March 31, 2020. Several vaccines have since been developed and deployed around the globe, helping to reduce disease severity and transmission of the virus.^1^ However, as of July 2021, infection rates were continuing to rise, an indication that the virus will be not be easily controlled. As a result, populations around the world are being forced to continue taking precautions to prevent COVID-19 infection.

The WHO has recommended avoiding “the 3Cs,” namely closed spaces, crowded places, and close contact with others.^2^ Physical distancing has formed a key part of recommendations by the WHO and governments around the world as a way to reduce the spread of COVID-19. The Japanese Ministry of Health, Labor, and Welfare, for example, has released practical examples of “new lifestyles,” recommending that people telework, work rotating shifts, keep their distance in the office, and conduct meetings online.^3^ However, such practices can lead to a greater likelihood of social isolation and loneliness. Thus, there is concern that continuing measures such as physical distancing to prevent COVID-19 may cause loneliness and have a negative impact on physical and mental health.^4^

A major problem that can accompany loneliness is increased alcohol consumption.^5^ Several studies have indeed reported an increase in alcohol consumption during the COVID-19 pandemic.^6–8^ Data from the United States shows that alcohol sales and deliveries increased after the COVID-19 outbreak.^9^ In Belgium, there was a 30.3% increase in alcohol consumption,^10^ and 21% of Canadians who were unlikely to go out drinking before the pandemic have started to do so.^11^ Similar to before the pandemic, negative psychological stress, anxiety, and loneliness are suggested causes of the increase in alcohol consumption during the COVID-19 pandemic. In addition to loneliness, social isolation is also associated with mental health problems.^12,13^ While loneliness and social isolation are similar, they differ in that the former is subjective while the latter is objective.^14^ Social isolation is thought to be a predictor of loneliness, which subsequently leads to psychological problems such as depression and anxiety.^15^

COVID-19 prevention measures like telework are leading to more social isolation and loneliness among the working population. This study aimed to assess whether loneliness among workers in Japan during the COVID-19 pandemic is associated with changes in alcohol consumption.

## Methods

### Study Design and Subjects

This investigation formed part of the Collaborative Online Research on the Novel-Coronavirus and Work (CoroNaWork) Project, a cross-sectional study that used internet surveys between December 22 and 26, 2020 to examine the health of Japanese workers during the COVID-19 pandemic. A detailed description of the protocol is published elsewhere.^16^ In brief, workers who indicated they were employed during the survey period were selected according to their prefecture of residence, occupation and sex. Workers’ questionnaire data were collected. Those with extremely short response times, height less than 140 cm, weight less than 30 kg, and inconsistent responses to multiple identical questions were deemed to have provided fraudulent responses and were excluded. Of the 33302 workers who returned the questionnaire, 6266 were excluded for providing fraudulent responses, leaving 27036 for analysis.

This study was conducted with the approval of the Ethics Committee of the University of Occupational and Environmental Health, Japan (Approval number R2-079). Informed consent was obtained through the questionnaire website.

### Assessment of loneliness and social isolation

Loneliness was assessed using the question: “During the last 30 days, how frequently did you feel the following emotions?” Participants responded by choosing from never, a little, sometimes, usually, and always. For analysis, responses of “never” and “a little” were categorized as indicating no loneliness, while responses of “sometimes,” “usually,” and “always” were categorized as indicating loneliness.

Social isolation was assessed using three questions about whether the participants had friends to talk to, acquaintances to ask for favors, and people to communicate with through social networking sites. Participants chose from yes and no responses.

### Assessment of usual and changes in alcohol consumption

A questionnaire was used to assess usual alcohol consumption and whether that consumption had changed during the COVID-19 pandemic. Drinking frequency was classified as at least two days per week, one day per week, or hardly ever. For analysis, alcohol consumption during the COVID-19 pandemic was classified as either increased or not increased.

### Other covariates

The following were included as confounders: age, sex, marital status, equivalent income, education, smoking, job type, number of employees at the workplace, cumulative incidence rate of COVID-19 in the prefecture of residence, lack of friends to talk to, lack of acquaintances to ask for favors, and lack of people to communicate with through social networking sites. Age was treated as a continuous variable, while all other variables were used as categorical variables and presented as percentages.

Further, the cumulative incidence of COVID-19 in the prefecture of residence in the month immediately preceeding the survey was treated as a community-level variable. These incidence data were obtained from the websites of public institutions.

### Statistical analysis

Univariate and multivariate logistic regression analyses were used to determine odds ratios (ORs), with loneliness or drinking frequency as independent variables and change in alcohol consumption as a dependent variable. The multivariate model was adjusted for age, sex, marital status, equivalent income, education, smoking, alcohol consumption, job type, number of employees at the workplace, cumulative incidence rate of COVID-19 in the prefecture of residence, lack of friends to talk to, lack of acquaintances to ask for favors, and lack of people to communicate with through social networking sites. A p-value less than 0.05 indicated statistical significance. Stata (Stata Statistical Software: Release 16. College Station, TX: StataCorp LLC.) was used for analysis.

## Results

Table 1 summarizes participants’ general characteristics. A total of 2831 of 27036 participants (10.5%) indicated they had increased their alcohol consumption during the COVID-19 pandemic. While there were no differences in age, marital status, education, or company size between those who did and did not increase their alcohol consumption, those who did so tended to be male, smokers and have higher income. There were no missing data among the responses of the 27036 people included in the analysis.

Table 2 summarizes the ORs of loneliness associated with changes in alcohol consumption according to the logistic model. The age-adjusted model revealed a significant association between increased alcohol consumption and loneliness (OR=1.78, 95%CI 1.59–1.98), as did the multivariate analysis (OR=1.94, 95%CI 1.70–2.21). There was also a significant association between increasing alcohol consumption and a lack of friends to talk to in both the age-adjusted model (OR=1.10 95%CI 1.02–1.20) and multivariate analysis (OR=1.14 95%CI 1.01–1.29). While a lack of acquaintances to ask for favors was also associated with increasing alcohol consumption in the age-adjusted model (OR=1.16 95%CI 1.07–1.26) and multivariate analysis (OR=1.22 95%CI 1.09–1.38), a lack of people to communicate with through social networking sites was significantly associated with no increase in alcohol consumption in both the age-adjusted model (OR=0.89 95%CI 0.82–0.97) and multivariate analysis (OR=0.85 95%CI 0.77–0.94).

Compared to those with no drinking habit, both those who drank less than one day per week and those who drank two or more days per week were significantly more likely to increase their alcohol consumption. Since the interaction term was significant, we further estimated the OR of loneliness associated with increased alcohol consumption for each category of drinking frequency using a logistic model. The results are shown in Table 3. Among those who were drinking two or more days per week, increasing alcohol consumption was significantly associated with loneliness in the age-adjusted model (OR=2.15 95%CI 1.87–2.47) and multivariate analysis (OR=1.98 95%CI 1.71–2.30). A similar finding was observed for those who drank less than one day per week: increasing alcohol consumption was significantly associated with loneliness in the age-adjusted model (OR=1.91 95%CI 1.40–2.58) and multivariate analysis (OR=1.51 95%CI 0.71–3.25). In contrast, among those who hardly ever drank, increased drinking was not associated with loneliness either in the age-adjusted model (OR=1.51 95%CI 0.71–3.25) or multivariate analysis (OR=1.22 95%CI 0.55–2.72).

## Discussion

### Main findings of this study

Approximately 10% of workers examined in this study increased their alcohol consumption during the COVID-19 pandemic. Loneliness, lack of friends to talk to, and lack of acquaintances to ask for favors were all factors linked to increased drinking habits. Meanwhile, a lack of people to communicate with through social networking sites was linked to no increase in alcohol consumption. The present study also found that the association between loneliness and increased alcohol consumption varied depending on participants’ prior drinking habits: those with a greater prior drinking habit demonstrated a greater association between loneliness and increased drinking.

### What is already known on this topic

Alcohol consumption has been reported to increase during natural disasters and infectious disease outbreaks. For example, reports indicate that people tended to drink more during the 2004 Indian Ocean tsunami and Hurricanes Katrina and Rita in 2005 than before the disasters.^17,18^ In addition, alcohol consumption increased during the Severe Acute Respiratory Syndrome (SARS) pandemic.^19,20^ Negative coping behaviors like drinking are used to suppress loneliness, depression and anxiety.^21^

Previous studies have suggested that whether or not alcohol consumption increases during the COVID-19 pandemic depends on a person’s usual alcohol consumption.^22–24^ Heavy and frequent drinkers tend to increase their alcohol consumption, while occasional drinkers show no change in their drinking habit.

While loneliness and social isolation partially overlap, they have important differences.^14^ People who are socially isolated do not necessarily experience loneliness, but, rather, can feel comfortable with their situation. An example of this is the “super solo” phenomenon in Japan.^25^ Super solo culture considers social isolation an advantage and those who engage in it enjoy being isolated without feeling lonely.

Social media can be useful for reducing loneliness while social distancing during the COVID-19 pandemic because it provides a means of communication with others.^26,27^ However, due to the overflow of inaccurate information, among other factors, social media has been linked to deterioration in mental health during the COVID-19 pandemic.^4^

### What this study adds

This study showed that an increase in alcohol consumption was associated with loneliness during the COVID-19 pandemic. Although previous studies have shown that COVID-19 has increased loneliness and that loneliness is associated with increased alcohol consumption.^4,7^ In this study, we link loneliness to increased drinking in the COVID-19 pandemic. Our findings suggest that loneliness, even among occasional drinkers, may increase both drinking and the risk of inappropriate drinking habits.

In addition to loneliness, we also examined social isolation based on whether participants had friends to talk to, acquaintances to ask for favors, and people to communicate with through social networking sites. Social isolation was defined as having a lack of friends to talk to and acquaintances to ask for favors; these factors, in addition to loneliness, were associated with increased alcohol consumption. In contrast, a lack of people to communicate with through social networking sites, which was also defined as social isolation, was associated with no increase in alcohol consumption. Thus, we speculate that use of social networking sites likely does not relieve feelings of loneliness, but may actually increase negative emotions like anxiety, which are linked to increased alcohol consumption.

### Limitations of this study

This study had several limitations. First, because this study was conducted using Internet monitors, generalizability of the results is unclear. However, we attempted to minimize subject bias as much as possible by sampling by region, occupation, and prefecture based on the incidence of infection. Second, we used a self-reported assessment of alcohol consumption in this study. Given that research has shown that drinkers tend to underreport their alcohol consumption,^28^ it is possible that the same occurred in this study. However, we think that underreporting was relatively unlikely because we used an anonymous survey conducted on the Internet. Third, we determined participants’ loneliness using one question based on previous studies.^29^ Further studies using other assessment methods are needed to confirm our results.^30,31^

## Conclusion

The COVID-19 pandemic has necessitated measures such as physical distancing, telecommuting, and mandates to refrain from going out and socializing. As a result, many people are now dealing with job and economic insecurity in addition to risk of infection. Against this backdrop are concerns about isolation and loneliness among workers and the related increase in alcohol consumption. We found that 10% of Japanese workers increased their alcohol consumption during the COVID-19 pandemic. For those with a drinking habit, increased drinking was associated with loneliness. Thus, education and measures against loneliness are needed to prevent inappropriate drinking habits among workers.

## Data Availability

All data produced in the present study are available upon reasonable request to the authors

## Author contributions

Y.F. was the chairperson of the study group. Y.K. conceived the research questions. All the authors designed the research protocol and developed the questionnaire. Y.K. conducted the statistical analysis with Y.F. Y.K. drafted the initial manuscript. All the authors revised and approved the final manuscript.

## Acknowledgements

The current members of the CORoNaWork Project, in alphabetical order, are Yoshihisa Fujino (present chairperson of the study group), Akira Ogami, Arisa Harada, Ayako Hino, Hajime Ando, Hisashi Eguchi, Kazunori Ikegami, Kei Tokutsu, Keiji Muramatsu, Koji Mori, Kosuke Mafune, Kyoko Kitagawa, Masako Nagata, Mayumi Tsuji, Ning Liu, Rie Tanaka, Ryutaro Matsugaki, Seiichiro Tateishi, Shinya Matsuda, Tomohiro Ishimaru, and Tomohisa Nagata. All members are affiliated with the University of Occupational and Environmental Health, Japan.

## Ethical approval

This study was approved by the ethics committee of the University of Occupational and Environmental Health, Japan (reference No. R2-079 and R3-006).

## Informed Consent

Informed consent was obtained in the form of the website.

## Data availability statement

The datasets for this study are available from the corresponding author on reasonable request.

